# The association between cancer and spousal rate of memory decline: a negative control study to evaluate (unmeasured) social confounding of the cancer-memory relationship

**DOI:** 10.1101/2020.02.24.20027516

**Authors:** Monica Ospina-Romero, Willa D. Brenowitz, M. Maria Glymour, Elizabeth R. Mayeda, Rebecca E. Graff, John S. Witte, Sarah Ackley, Kun Ping Lu, Lindsay C. Kobayashi

## Abstract

Cancer diagnoses are associated with better long-term memory in older adults, possibly reflecting a range of social confounders that increase cancer risk but improve memory. We used spouse’s memory as a negative control outcome to evaluate this possible confounding, since spouses share social characteristics and environments, and individuals’ cancers are unlikely to cause better memory among their spouses. We estimated the association of an individual’s incident cancer diagnosis (exposure) with their own (primary outcome) and their spouse’s (negative control outcome) memory decline in 3,601 couples from 1998-2014 in the Health and Retirement Study, using linear mixed-effects models. Incident cancer predicted better long-term memory for the diagnosed individual. We observed no association between an individual’s cancer diagnosis and rate of spousal memory decline. This negative control study suggests that the inverse association between incident cancer and rate of memory decline is unlikely to be attributable to social/behavioral factors shared between spouses.

---

Growing evidence consistently demonstrates an inverse relationship between cancer and Alzheimer’s disease (AD) and related dementias.^1,2^ This inverse relationship was observed in the population-based US Health and Retirement Study (HRS), where memory decline in individuals who developed cancer was slower than memory decline in those never diagnosed with cancer over follow-up; this difference emerged both prior to and after the cancer diagnosis.^3^ Cancer treatments, particularly chemotherapies, are neurotoxic, and acute memory decline has been observed immediately following diagnosis and treatment.^3,4^ Yet, the long-term inverse cancer-AD relationship is consistently reported, including in studies designed to mitigate biases that could explain the association, such as competing risks, diagnostic bias, and survival bias.^5–8^ The robustness of this relationship to a range of methodological approaches, as well as the unexpected favorable memory outcomes among individuals with cancer even before their diagnosis, indicate that the cancer and AD association is likely confounded by shared common causes.

Determining whether the inverse association between cancer and AD arises from common genetic or biological causes, as opposed to non-genetic factors, could provide insight into the biological mechanisms underlying carcinogenesis and neurodegeneration. Inverse genetic regulation of carcinogenesis and neurodegeneration has been postulated,^9^ while non-genetic factors such as socioeconomic, behavioral, and environmental factors (collectively, referred to here as ‘social factors’) also contribute to cancer and AD.^10^ There is limited evidence on the factors that could account for the cancer-AD association. One strategy to evaluate confounding is to use a negative control outcome that cannot be causally related to the exposure of interest, but is subject to similar confounding bias as the original association.^11^ In such a study, observing no association between the exposure and the negative control outcome indicates that the original association is unlikely to have arisen from unmeasured or residual confounding.^11^

Cancer and AD risk factors are commonly shared between spouses due to socially assortative partnerships, and spouses’ influence on each other’s behaviors, and social and physical environments.^12,13^ Having data on spousal memory function in the HRS provides an opportunity to evaluate unmeasured confounding by non-genetic social factors shared between spouses (as shown in Supplemental Figure 1). We conducted a negative control study to estimate the association between an individual’s cancer diagnosis (exposure) and their spouse’s rate of memory decline (negative control outcome). We assumed that genetic traits are negligibly correlated between spouses in the general population.^14^ We hypothesized that slower memory decline in individuals whose spouse developed cancer would suggest confounding by unmeasured non-genetic, social factors that are shared between spouses. An observation of null results would rule out meaningful confounding due to social factors, providing indirect evidence that genetic or biological factors may explain the inverse cancer-memory decline association.

## Methods

### Study design and sample

We followed the same study design as our previous study, which compared pre- and post-diagnosis rates of memory decline in adults with an incident cancer, to rates of aging-related memory decline in cancer-free adults in the US Health and Retirement Study (HRS).^3,15^ HRS respondents born before 1949 with interviews in 1998 (age 50+ at baseline), no history of cancer, and a co-residing spouse also in the HRS were eligible for this analysis (7202 individual respondents in 3601 couples; Supplemental Figure 1). All data were assessed in biennial interviews from 1998-2014 (up to 16 years of follow-up).

The HRS was approved by the University of Michigan Health Sciences Human Subjects Committee. These analyses were determined exempt from review by the University of California, San Francisco Institutional Review Board.

### Incident cancer

Incident cancer was assessed as self-reported physician diagnosis of cancer excluding non-melanoma skin cancer (1,212 respondents).

### Memory

Memory was assessed as immediate and delayed recall of a 10-word list. Memory scores were imputed from proxy assessments to retain severely impaired respondents in analyses.^16^ Memory scores were standardized using the baseline mean and standard deviation (SD) of the original sample.^3^ Time of cancer diagnosis was defined as time zero with respect to memory: memory assessments preceding each diagnosis were assigned negative time in years and memory assessments following diagnosis were assigned positive time in years. For the negative control analysis, the times of each spouse’s memory assessments were calculated with respect to the respondent’s diagnosis date. For respondents who did not report an incident cancer, time of cancer diagnosis was set to 0. In the cancer-free group, 1,528 respondents had shorter follow-up than their spouses. We carried forward their last cancer status observation to retain their spouse’s subsequent memory assessments in the negative control analysis.

### Covariates

Plausible confounders of the cancer-memory decline association were self-reported for each spouse: sex, race, childhood socioeconomic status^17^, years of education, baseline household wealth, self-rated childhood health, baseline vigorous physical activity, current smoking, alcohol use, body mass index, and prior diagnoses with hypertension, diabetes, heart disease, stroke, lung disease, or arthritis.

Supplemental Figure 1 depicts the hypothesized causal structure linking an individual’s cancer diagnosis to their rate of memory decline, with plausible measured and unmeasured confounders, and the spousal negative control outcome design.

### Statistical analysis

We examined the correlations between measured covariates within couples to determine likely shared confounders. Using multivariable linear mixed-effects models, we first replicated the previously observed association between an individual’s incident cancer and their own rate of memory decline^3^ (primary outcome) within the current study sample. Next, we swapped memory values at each time point between spousal pairs and re-ran the models to estimate the negative control outcome association between an individual’s cancer diagnosis and their spouse’s rate of memory decline. Individuals’ slopes of memory trajectories were modeled as random effects with random individual and household intercepts at 75 years of age. We included model terms for whether the respondent was diagnosed with cancer, timing of each memory assessment with respect to cancer diagnosis, and a separate time-dependent cancer indicator to account for acute change in memory function at the time of diagnosis. Models were adjusted for the respondents’ and their spouses’ measured confounders, consistent with Supplemental Figure 1.

Age at each interview and age at diagnosis (for individuals with cancer) was centered at 75 years, allowing for comparisons between predicted average memory function in individuals aged 75 immediately prior to diagnosis and predicted average memory function in cancer-free individuals aged 75. The supplemental methods contain detailed descriptions of the models. All analyses used Stata/SE version 15.1 (StataCorp).

## Results

Baseline characteristics and correlation coefficients for these characteristics within spouses are presented in Supplemental Table 1. The primary association between an individual’s cancer diagnosis and their own memory decline was successfully replicated (Figure 1A; Supplemental Table 1). Individuals diagnosed with cancer had better memory than cancer-free individuals immediately before diagnosis, and an acute memory decline at diagnosis. Long-term memory decline was slower in the cancer group both before and after diagnosis, compared to cancer-free individuals (Figure 1A, Supplemental Table 1). In the negative control outcome model (Figure 1B; Supplemental Table 1), spouses of incident cancer cases had no difference in memory function immediately before the diagnosis compared to spouses of individuals who never had a cancer diagnosis (0.032 SD units, 95% confidence interval (CI): −0.018, 0.082). An individual’s cancer diagnosis was not associated with acute change in their spouse’s memory at diagnosis (−0.001 SD units, 95% CI: −0.036, 0.033). Long-term rate of memory decline in spouses of individuals with a cancer diagnosis also did not differ before (difference: 0.022 SD units, 95% CI: −0.026, 0.069) or after (difference: 0.012, 95% CI: −0.040, 0.064) the diagnosis, compared to rate of memory decline in spouses of individuals without cancer (Figure 1B, Supplemental Table 1).

**Figure 1.**
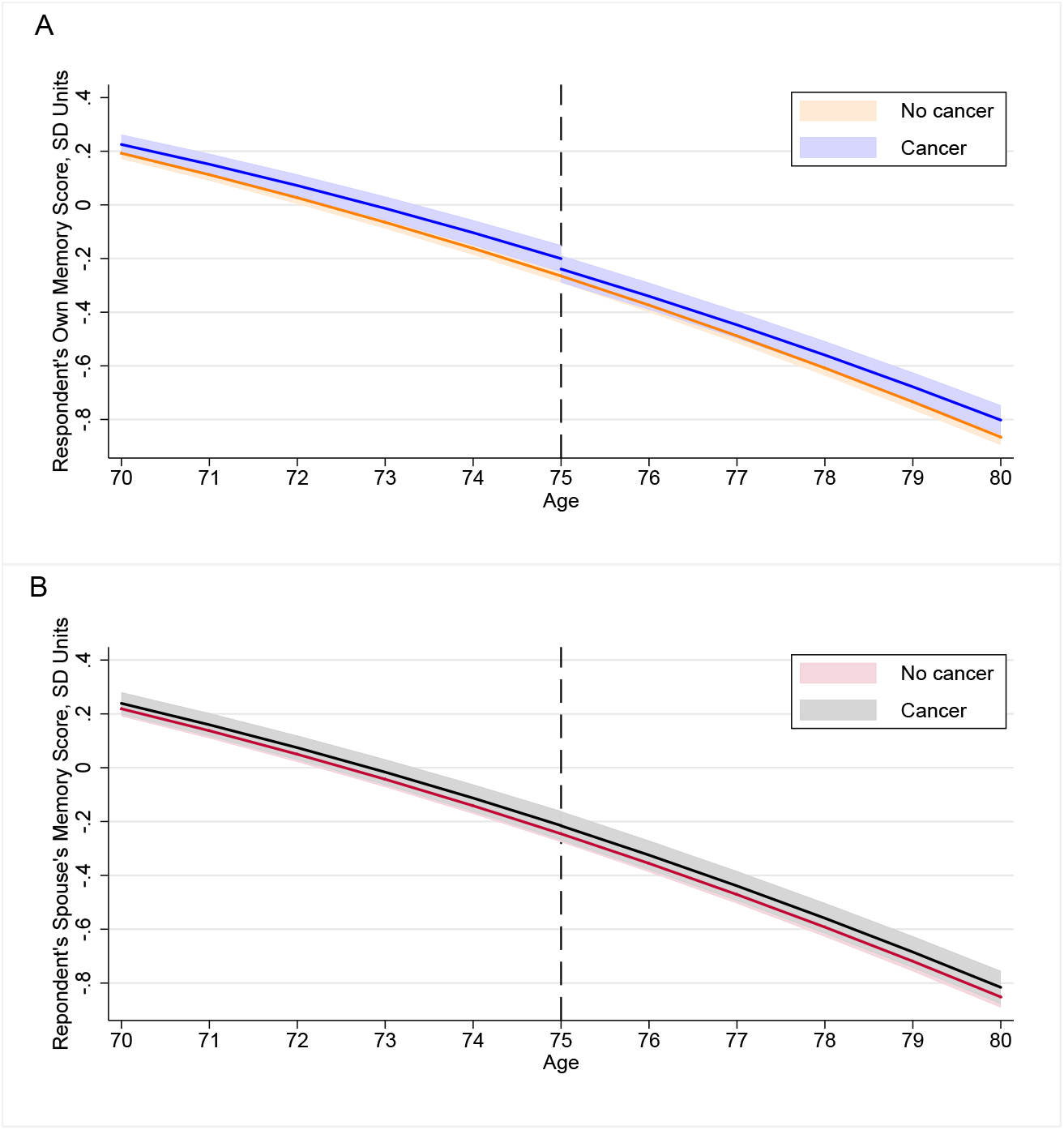
Predicted memory trajectories from linear mixed-effect models. **A**. Predicted memory trajectories and 95% CI (shaded area) for a person diagnosed with cancer at age 75 years (vertical line) in the reference categories (female, white, 12 years of education, no history of alcohol use or tobacco use, no baseline comorbidities), compared to memory trajectories in a person with the same characteristics, but with no cancer diagnosis during follow-up. **B**. Predicted memory trajectories in the spouse of a person diagnosed with cancer at age 75 years, compared to the spouse of a person with no cancer diagnosis during follow-up.

## Discussion

Our findings indicate that the previously observed inverse association between cancer and memory decline is not confounded by unmeasured, non-genetic factors shared between spouses. We employed the negative control study design because observational studies rarely capture the full range of socioeconomic, behavioral, and environmental conditions that could inversely affect cancer and AD risk, but these circumstances are commonly shared between spouses.^12,13,18^ Our findings are consistent with a growing body of evidence indicating that cancer and AD are inversely associated,^6,7^ and suggest that unmeasured biological or genetic factors might be driving this association.

Limitations are similar to those of the original study, and include survival bias due to differential follow-up times (although this is less likely with multiple assessments of a continuous outcome) and lack of data on non-memory cognitive domains (although memory decline is the hallmark of AD^19^). The relatively small sample of spouses resulted in some imprecision in our effect estimates. The negative control analyses assumes that measurement error is equivalent between the original and negative control outcomes—a reasonable assumption for memory function measures in this cohort of spouses.^20^ Although spouses may experience an acute decrease in cognition after cancer diagnosis due to stress, depression, or caregiving burden, we did not observe this outcome in our data.^21^

In summary, these findings suggest a common biological or genetic cause acting in opposite directions in carcinogenesis and neurodegeneration. Improved understanding of the potential shared biological mechanisms of cancer and AD may result in novel preventive and therapeutic strategies for both conditions.

## Data Availability

Drs Ospina-Romero and Kobayashi had full access to all the data in the study and take responsibility for the integrity of the data and the accuracy of the data analysis. The Health and Retirement Study is publicly available database. Access to this data should be requested to the University of Michigan.

https://hrs.isr.umich.edu/welcome-health-and-retirement-study

## Abbreviations

AD: Alzheimer’s disease
HRS: Health and Retirement Study
SD: standard deviation
CI: confidence interval.

## Acknowledgements

This study was supported by grant R01AG059872 (Glymour, Graff, Mayeda, Ospina-Romero, Witte) and grant K01AG062722 (Brenowitz) from the National Institute on Aging, grant AARF-18-565846 (Brenowitz) from the Alzheimer’s Association, and grant R03CA241841 (Kobayashi) from the National Cancer Institute.

---

**Table 1.**
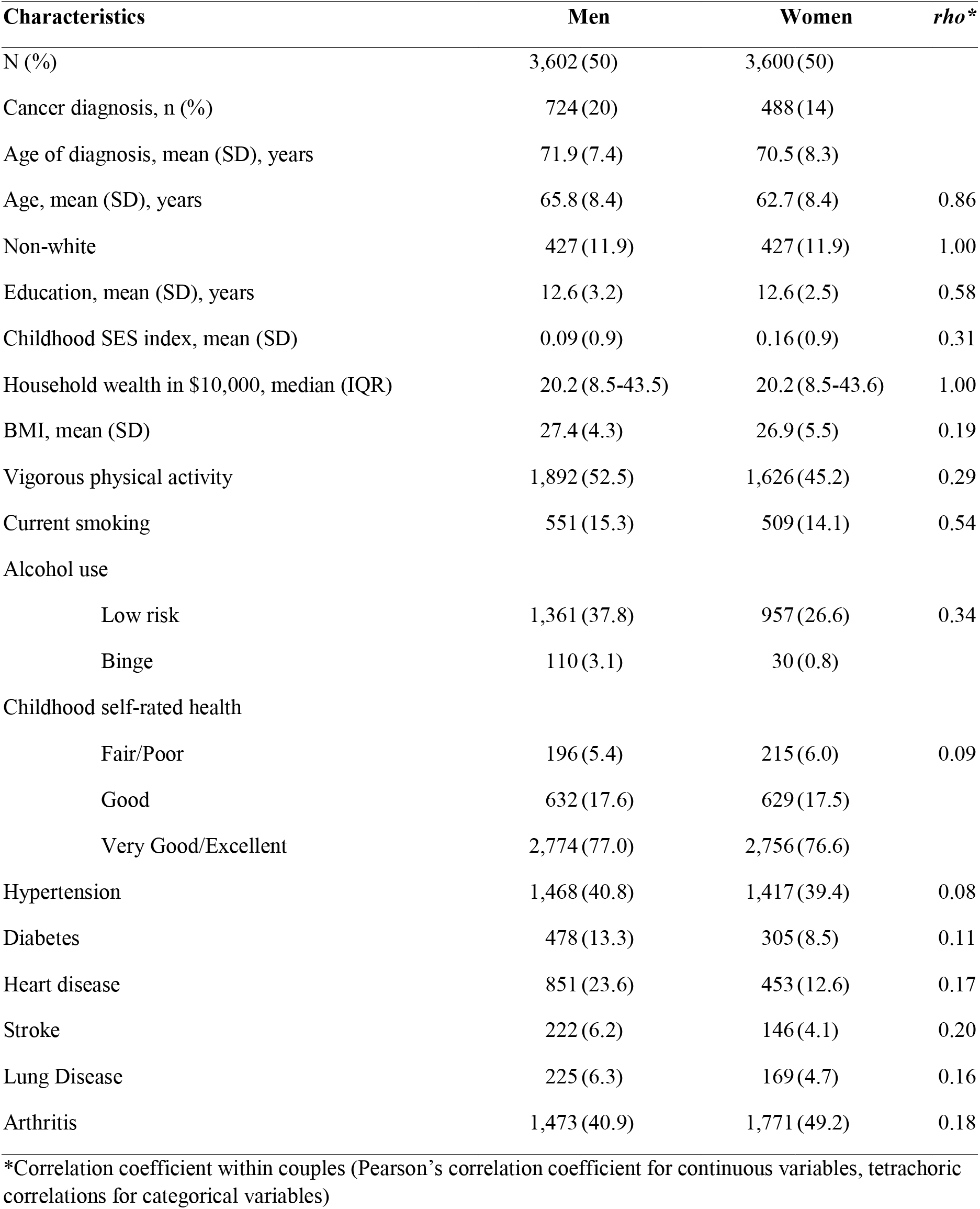
Baseline Characteristics of Couples Who Were Cancer-Free in 1998, US Health and Retirement Study, United States.

## Notes

### Competing Interest Statement

The authors have declared no competing interest.

### Clinical Trial

Not a clinical trial

